# Laboratory Comparison of Rapid Antigen Diagnostic Tests for Lymphatic Filariasis: STANDARD^TM^ Q Filariasis Antigen Test (QFAT) and Bioline Filariasis Test Strip (FTS)

**DOI:** 10.1101/2024.03.25.24304827

**Authors:** Patricia M Graves, Jessica L Scott, Alvaro Berg Soto, Antin YN Widi, Maxine Whittaker, Derek Lee, Colleen L Lau, Kimberly Y Won

## Abstract

**Background:** Accurate and user-friendly rapid diagnostic tests (RDT) are needed to assess prevalence of *Wuchereria bancrofti* antigen in the Global Programme to Eliminate Lymphatic Filariasis (GPELF). We evaluated performance under laboratory conditions of the new Q Filariasis Antigen Test (QFAT) against the Filariasis Test Strip (FTS) for detecting antigen of *Wuchereria bancrofti*, a causative agent of lymphatic filariasis (LF).

Methodology/Principal Findings: We compared test performance using available panels of serum (n=195) and plasma (n=189) from LF endemic areas in the Asia-Pacific region (Samoa, American Samoa and Myanmar) together with Australian negative controls (n=46). Prior antigen and antibody positivity status of endemic samples had been determined by rapid test or ELISA. The proportion of all samples testing positive at 10 minutes was higher with QFAT (44.8%) than FTS (41.3%).

Concordance between tests was 93.5% (kappa 0.87, N=417) at 10 minutes, and increased over time to 98.8% (kappa 0.98) at 24 hours. Sensitivity of QFAT and FTS at 10 minutes compared to prior antigen results were 92% (95% CI 88.0-96.0) and 86% (95% CI 80.0-90.0), respectively. Specificity was 98% for QFAT and 99% for FTS at 10 minutes. Sensitivity increased over time for both tests, rising to 99% for QFAT and 97% for FTS at 24 hours. QFAT positively identified all microfilaria (Mf)-positive samples, whereas FTS was negative for 3 of 66 Mf-positives. For both QFAT and FTS, there was evidence of cross-reaction with *Dirofilaria repens* and *Onchocerca lupi* but not with *Acanthochilonema reconditum*, *Cercopithifilaria bainae or Strongyloides.* Disadvantages noted for QFAT were inconvenient packaging and an additional buffer step. Advantages of QFAT include ease-of-use, smaller sample (10-20 µL vs 75 µL for FTS), clearer control line, and higher sensitivity for Mf-positive samples.

Conclusions/Significance. Under lab conditions, QFAT is a suitable rapid Ag test for use in filariasis elimination programmes and has advantages over FTS.

**Author summary:** Lymphatic filariasis (LF) is a debilitating and stigmatizing disease that affects populations in tropical areas usually in developing social environments. It is caused by a parasite worm transmitted by mosquitoes. The WHO programme to eliminate LF aims to improve the lives of their residents through a global mass drug administration campaign, and provide the tools to monitor prevalence within the countries’ public health contexts. It is imperative to utilize the most effective and practical diagnostic tests to monitor progress to elimination of this disease, while ensuring a cost-effective and rapid implementation under potentially vulnerable settings. In this study we investigated the performance of a new rapid antigen diagnostic test for LF compared to the existing recommended test, in samples of known infection status from the Asia-Pacific region. The results showed the new test to be a suitable rapid antigen test, with advantages over the current test, for use in filariasis elimination programmes in the region.

## Introduction

Lymphatic filariasis (LF) is a mosquito-transmitted neglected tropical disease caused by infection with filarial parasites *(Wuchereria bancrofti*, *Brugia malayi*, or *B. timori*). Over time, infection can lead to damage of the lymphatic vessels, causing hydrocele and lymphoedema. People who live with these chronic and disabling manifestations of LF can experience reduced economic productivity and social stigma. The Global Programme to Eliminate Lymphatic Filariasis (GPELF), established by the World Health Organization (WHO), aims to eliminate LF as a public health problem with a two-armed approach: interrupting transmission through mass drug administration (MDA) of anti-filarial medicines and alleviating suffering among patients through morbidity management and disability prevention.

Currently, 44 of 72 LF endemic countries still need MDA, with the majority implementing it with support from GPELF [1]. There has been a 74% reduction in global LF infections from 1997 to 2018 [2] but many countries are still conducting MDA or surveillance to validate elimination.

Accurate and sensitive diagnostic tests for measuring infection prevalence are essential components of the GPELF [3, 4]. National programs use diagnostic tests to establish baseline endemicity, monitor the impact of MDA campaigns, inform decisions to stop MDA in transmission assessment surveys (TAS), and detect potential recrudescent transmission post-cessation of MDA. Historically, the gold standard test for diagnosing LF was identifying microfilariae (Mf) on stained blood slides, but this requires time and skill, and (outside the Pacific areas of subperiodic diurnal transmission) blood samples for slides must be collected at night. In the 1980s, two enzyme-linked immunosorbent assays (ELISA) were developed to detect *W. bancrofti* using monoclonal antibodies to detect the same circulating filarial antigen (Ag) from adult worms [5-7]. Both tests produce quantitative readouts and can be performed using samples derived from either dried blood spots, anticoagulated whole blood, serum or plasma, with attention to dilution factors [8]. However, these tests require a suitably equipped laboratory.

One of the Ag ELISA tests, Og4C3, is commercially available (Cellabs, Sydney). The other ELISA test [7] was subsequently converted to a rapid test format, the immunochromatographic test (ICT) (initially Binax Now, then Alere, now Abbott) [9]. The ICT was widely utilised since the start of the GPELF in national surveys for prevalence mapping and surveillance. Subsequently, to reduce the cost and amount of blood needed for ICT, and improve the shelf life and storage requirements, the Alere filariasis test strip (FTS) was developed using the same critical reagents in a new test format [10]. Despite some discrepancies in concordance between the ICT and FTS tests in field evaluations [11], with FTS showing increased sensitivity, the WHO Neglected Tropical Diseases Strategic and Technical Advisory Group was satisfied with the diagnostic characteristics of FTS compared with ICT [12]. Thus, guidance on implementing TAS and critical cut-off numbers for passing were not changed and a study in American Samoa supported the approval of the FTS test for use in programme monitoring [11]. Since 2015, where *W. bancrofti* is the causative agent of LF, the Bioline Filariasis Test Strip (FTS, Abbott) has been the main Ag rapid diagnostic test recommended for programme use.

While the FTS has been used widely since 2015, limitations have been identified. The assay cross-reacts with *Loa loa* infections [13, 14] although this is not a concern in the Asia-Pacific region. The test requires a relatively large volume of blood (75 µL), routinely collected by finger prick. The time needed to collect 75 µL increases the risk of clotting and could prevent proper flow of blood through the test strip. Logistical challenges have been experienced in field settings because the FTS consists of a single lightweight strip devoid of any protective housing. Users are required to secure the test strips onto a plastic tray with labels or tape to minimize movement. Issues with flow of whole blood through the strip has caused some users to delay the start of the 10-minute timing for reading the test result. Therefore, improved and more user-friendly rapid antigen tests are urgently needed. Availability of alternative tests would also ease supply problems with FTS such as those experienced during the COVID-19 pandemic when all rapid test manufacturers were focused on producing rapid antigen tests for SARS-CoV-2. In addition, FTS is thought to be less accurate when used with EDTA than heparinized blood samples.

Although LF rapid Ag tests and similar rapid antibody (Ab) tests are designed primarily to give a binary (positive/negative) result, they can also give a semi-quantitative readout by comparing the density of the test line to the control line. Intensity is usually scored on a scale of 1 to 3, where 1 (low) represents a test line less dense than control, 2 (medium) when the two lines are of similar density, and 3 (high) when the test line is denser than the control line. Changes in Ag or Ab intensity score over time have been used to assess impact of control measures and progress to elimination for LF [15] and onchocerciasis [16]. Chesnais et al., 2016 reported a strong correlation (ρ = 0.91; *P* < 0.001) between the FTS intensity score and plasma Ag levels measured by Og4C3 ELISA in a study in Democratic Republic of the Congo in 2014, supporting the use of test line density as a proxy for blood Ag level. Higher test density scores were also associated with Mf slide positivity [15]. Semi-quantitative intensity scoring is frequently done in LF surveys, but its usefulness in cross-sectional studies is not yet clear.

The primary aim of this study was to evaluate the diagnostic performance of QFAT compared to the currently recommended FTS through a head-to-head comparison of samples with known prior Ag results. The WHO Collaborating Centre for Vector-borne and Neglected Tropical Diseases at James Cook University (JCU) served as an independent assessor for this study. We used samples collected from the Asia-Pacific region that were already available in JCU collections. Results will inform the decision about whether the new Standard Q Filariasis Antigen Test (QFAT) can serve as a suitable alternative to the FTS in GPELF activities. Secondary objectives were to investigate cross-reactivity with other helminths, discordance in results between independent test readers, and performance of the tests with different sample types (serum, heparin plasma, EDTA plasma). In addition, we studied whether test line density was informative, and whether there were changes in test results (concordance, sensitivity and specificity) over time. The requirement to read all tests at 10 minutes can be challenging if many samples are tested concurrently during large surveys. Extending the reading time without compromising specificity would help to better distribute the workload, especially for duplicate readers, and allow later checking by supervisors.

## Materials and Methods

### QFAT and FTS Kits

The WHO provided 510 FTS tests (17 boxes of 30 tests) and SD Biosensor provided 500 QFAT tests (20 boxes of 25 tests). Tests were sent to JCU Cairns and kept at room temperature (∼25℃) for up to 3 months before use. The batch numbers were FTS lot number 181193; FTS pouch number 178066; QFAT lot number SIJ35HIAC and QFAT buffer number 5IJ34D1S5. Expiry dates were 28 Dec 2022 for FTS and 16 Jan 2024 for QFAT. The study was performed in August and September 2022. Initially one of each test type was checked with positive control obtained in 2014 from the Filariasis Research Reagent Resource Center (FR3) http://www.filariasiscenter.org/; both tests gave a weak positive result (as expected). This was repeated with one of each test type at the end of the study, with similar results.

Both FTS and QFAT are designed to be used with whole blood direct from a participant’s finger. For FTS, it is generally accepted that if anticoagulated blood is used, heparin should be the choice of anticoagulant. QFAT also gives instructions for use of the test with serum or plasma, with a reduced sample volume compared to whole blood.

### Blood sample description and selection

A total of 456 serum and plasma samples were assembled from collections archived at JCU and affiliated research groups. Samples included those previously tested for LF antigen by ICT, FTS or Og4C3 ELISA (and in some cases, also for Mf and antibodies) in previous studies in American Samoa, Samoa and Myanmar [17-19, 11]. These samples were collected either from LF endemic areas (n=384) or from non-LF endemic countries (n=72) for control purposes. The 384 endemic samples were collected from American Samoa (total n=257: 133 collected in 2014 and 124 in 2016), Samoa (n=35 in 2019) and Myanmar (n=92 in 2014). Samples of EDTA plasma were included to investigate whether QFAT would be a more versatile test than FTS for different sample types. The 72 non-endemic samples were mainly from Australia (46 human negative controls and 19 human samples positive for *Strongyloides* antibody diagnosed at NSW Pathology by ELISA against *S.ratti* antigen) together with seven dog samples from Italy used in a previous study of Og4C3 ELISA [20]. Of the 384 endemic area samples, LF antibody results were available for only 349 (Supplementary Table 1). Slide Mf results were only available from American Samoa and Samoa for 165 (78.2%) of the Ag-positive participants, of whom 69 were Mf-positive.

It should be noted that American Samoa 2016 samples of serum (collected in SST vacutainers) and plasma (collected in heparin tubes) were from the same individuals (N=62). In Samoa 2019, there were 12 individuals who had plasma collected in both heparin and EDTA tubes. Details of sample collections are available in previous publications [17-19, 21].

Two of the 456 samples (one heparin and one EDTA plasma) had no serum or plasma in selected vial (missing) and were removed from study entirely. Two additional samples were unable to be tested using FTS due to insufficient volume (one heparin and one EDTA plasma). Additionally, 18 out of 19 of the human *Strongyloides* positive samples and one of the Australian negative controls only had enough sample for testing by QFAT, not FTS. Samples selected and tested are shown in Figure 1.

**Fig 1.**
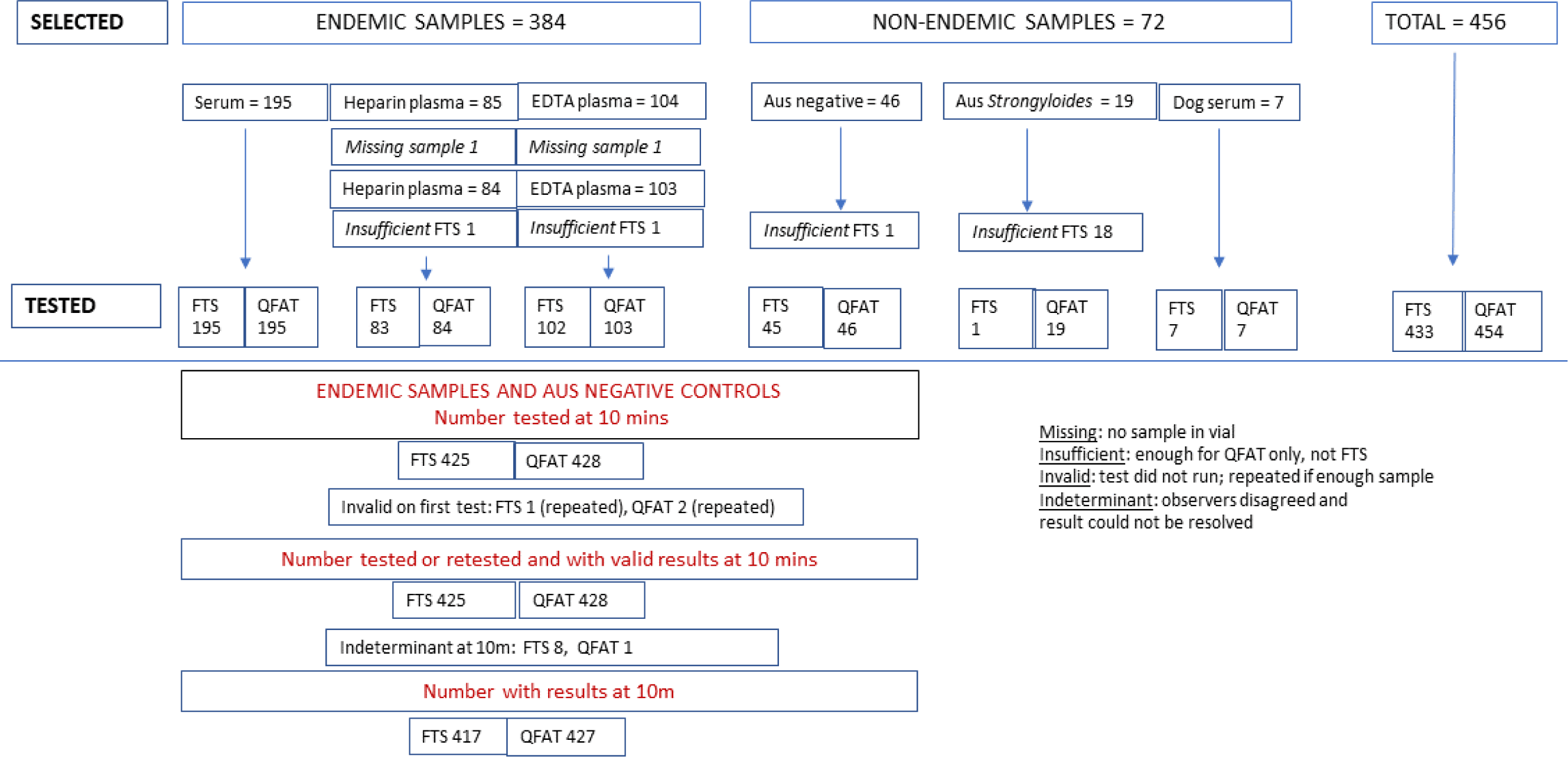
Flowchart of samples selected, tested and indeterminant at the 10 minute reading, by sample category and type.

Samples were selected with the aim of having approximately equal proportions of Ag-positive and Ag-negative samples for optimal estimation of sensitivity and specificity. The final selection resulted in 54.9% Ag-positives and 45.1% Ag-negatives among the samples from endemic areas (Supplementary Table 1).

### Ethical approval

Participants gave express written consent for their samples to be stored and used for additional studies, with the exception of Myanmar where waiver of consent for re-use of deidentified samples was applied for. Samples were collected under the following ethical approvals:

- American Samoa 2014: Institutional Review Board of American Samoa, James Cook University Human Research Ethics Committee and The University of Queensland (approval number 2014000409).
- American Samoa 2016: Australian National University (protocol number 2016/482)
- Samoa 2019: Samoan Ministry of Health and The Australian National University Human Research Ethics Committee (protocol number 2018/341).
- Myanmar 2014: James Cook University Human Research Ethics Committee approval number H5261 approved by the Ministry of Health and Sports, Myanmar. Since consent in the initial study for future sample use was not fully explicit and participants could not be recontacted, waiver of consent was granted by James Cook University Human Research Ethics Committee (approval number H8341) in consultation with Myanmar collaborators.

### Sample preparation and test procedures

The samples used for testing had been shipped frozen from the country of origin to Australia and stored at minus 70°C for three to six years depending on the source. Human control samples had been stored for varying periods from six to over 10 years. Most samples had undergone one previous freeze-thaw cycle. They were thawed from minus 70°C, aliquoted and kept at 4°C until testing the same day. QFAT and FTS tests were performed as per the manufacturers’ instructions, but instead of using the pipettes supplied by the kits, calibrated micropipettes were used to transfer the appropriate sample volumes to the test sample pads.

The results of QFAT and FTS were read at the recommended time of 10 minutes by two independent blinded observers who were unaware of the sample type or prior result. Each observer used the manufacturers’ criteria for test interpretation and classified the tests as positive, negative or invalid. Tests were deemed invalid if no control line was present in the test window. A 10-minute timer was started from the time that the sample migrated to the test line (FTS) or time of adding buffer (QFAT).

For FTS and QFAT tests that were Ag-positive, the intensity of the test line relative to the control line was scored as low (test line intensity lighter than the control line), medium (test line intensity same as control line) or high (test line intensity darker than the control line). At 1 hour and 24-hour time points after initial testing, QFAT and FTS tests were re-read by the same two observers. Flashlights were used for illumination to assess test lines if needed.

Recommended volume for whole blood for the FTS test is 75 µL. Current instructions do not specify a volume for serum or plasma. Recommended volume for whole blood for the QFAT test is 20 µL, with 10 µL recommended for serum. Initial testing of eight samples of Ag-positive serum and heparin plasma from American Samoa (4 each of Mf positive and negative) was conducted to establish sample volume for FTS for this study and the integrity of the tests. Ideally, we would have compared the same volume on both tests (for example 20 µL). However, with FTS, none of eight serum samples flowed with 20 µL, giving invalid results, while the QFAT test functioned well with 10 or 20 µL. Based on these preliminary evaluations, we used 75µL for FTS and 10 µL for QFAT (manufacturers’ recommendations) for the rest of the samples.

### Data analysis

Results were recorded on paper, transcribed into Excel and imported into R studio (v. 2023.06.0 Build 421) for recoding and analysis. Records with missing or inconsistent information were identified and rechecked against paper records. Selected samples were classified as missing (no serum or plasma in selected vial), or insufficient if they had not enough volume for testing.

#### Test validity

The proportion of invalid tests was based on the initial 10-minute reading to approximate the real-world situation. However, tests deemed invalid at the first 10-minute reading were repeated if there was sufficient sample remaining, to maximise the sample size for concordance and performance analysis.

#### Discordance between observers

The proportion of tests with discordance between the two observers was determined. Results were stratified by whether the samples originated from an endemic or non-endemic region and then by sample type (i.e., endemic serum, endemic heparin plasma, endemic EDTA plasma, non-endemic human serum, and non-endemic dog serum). McNemar’s Chi-squared test was performed to assess the difference between FTS and QFAT for discordant results for each sample type. Only samples with valid test results for both FTS and QFAT were included in this analysis.

#### Antigen prevalence

For valid tests, the following rules were used to assign a final result to each sample (positive, negative, or indeterminant) at each time point:

a. If both observers agreed on positive or negative, the result was assigned as positive or negative, respectively.
b. If the observers disagreed and a third assessment was made on a repeat test with one observer to break the tie, the dominant result was assigned (e.g., a positive result if 2 out of 3 assessments were positive).
c. If the observers disagreed and a third assessment (repeat test) was not able to be conducted, the result was classified as indeterminant.

For assessment of concordance and performance, *Strongyloides*-positive human samples and dog serum samples were excluded, leaving a sample size of 430 comprising endemic samples and Australian negative controls. Missing, insufficient and indeterminant results were also excluded for analysis at each of the three reading times.

#### Concordance between test types

To determine concordance between FTS and QFAT results, we calculated the percentage of agreement between positive and negative results and the Cohen’s Kappa (K) agreement statistic with 95% CI for each of the three reading times. Classification of K was as follows: poor (K <0.2), fair (K 0.21-0.40), moderate (K 0.41-0.60), good (K 0.61-0.80) and excellent (K 0.81-1.00). Kappa values were obtained using the VCD package for R, version 1.4-11. Only records with valid results for both FTS and QFAT were included.

#### Test performance

We compared the final result for each sample in each test to their results obtained in previous studies (Supplementary Table 1). LF Ag and Ab testing was not previously performed on all the Australian negative samples included in this analysis, although some had been used in previous ELISA studies as negative controls. Considering that LF has not been endemic in Australia for almost seven decades, we assumed that these samples were negative for LF Ag and Ab. We compared results in this study with two reference standards from previous testing: 1) the prior Ag test result, and 2) the prior combined ‘Ag or any Ab’ result. We calculated the sensitivity, specificity, positive and negative predictive value of FTS and QFAT against each of these reference standards using epiR package v 2.0.62. This analysis was performed at all three time points to assess whether test performance metrics changed over time.

Results for FTS and QFAT were also compared to Mf-positivity results from previous studies. In these studies, slides were made from Ag-positive persons only.

#### Intensity score comparison, by observer

We used the entire dataset (n = 452 samples for FTS and 454 samples for QFAT) to classify samples by their intensity score value (either low, medium or high) at each time point (i.e., 10 mins, 1 hour and 24 hours), stratified by observer and test type. Samples missing values for at least one time reading were also removed.

#### Test usability

At the end of the study, the two test observers independently provided written feedback on each test, prompted by the following categories: Instructions for use, test packaging, test set-up, sample volume, control line, test readability, and other comments.

## Results

A total of 456 samples were selected for testing, of which 384 were from endemic areas; two endemic samples were missing for both tests (no serum or plasma in selected vial).

### Test validity

At the first 10-minute reading, one of the FTS tests (0.2%, N=433) and two of the QFAT tests (0.4%, N=454) gave invalid results. All three samples were repeated, providing valid results for 380 FTS samples and 382 QFAT samples from endemic areas. None of the non-endemic samples tested (N= 53 FTS and 72 QFAT) had invalid results.

### Discordance between observers

Discordance between the two observers at 10 minutes with endemic samples was greater for FTS than QFAT readings (3.7% versus 1.8% respectively, p=0.096). Discordance in endemic samples was greatest for EDTA plasma samples using FTS (6.8%) and lowest for serum samples using QFAT (1.0%). These differences were not statistically significant at the p<0.05 level (Supplementary Table 2). There were no discordant observations in non-endemic samples for either FTS or QFAT. Discordant results at any time point that could not be repeated were classed as indeterminant, with details given in Supplementary Fig 1.

### Overall positivity, by test type

Counts of positive, negative, and indeterminant endemic samples by sample type and test type at the 10-minute reading are shown in Table 1A. In general, QFAT provided more positive results than FTS for every sample type, as well as producing fewer indeterminant results.

**Table 1:**
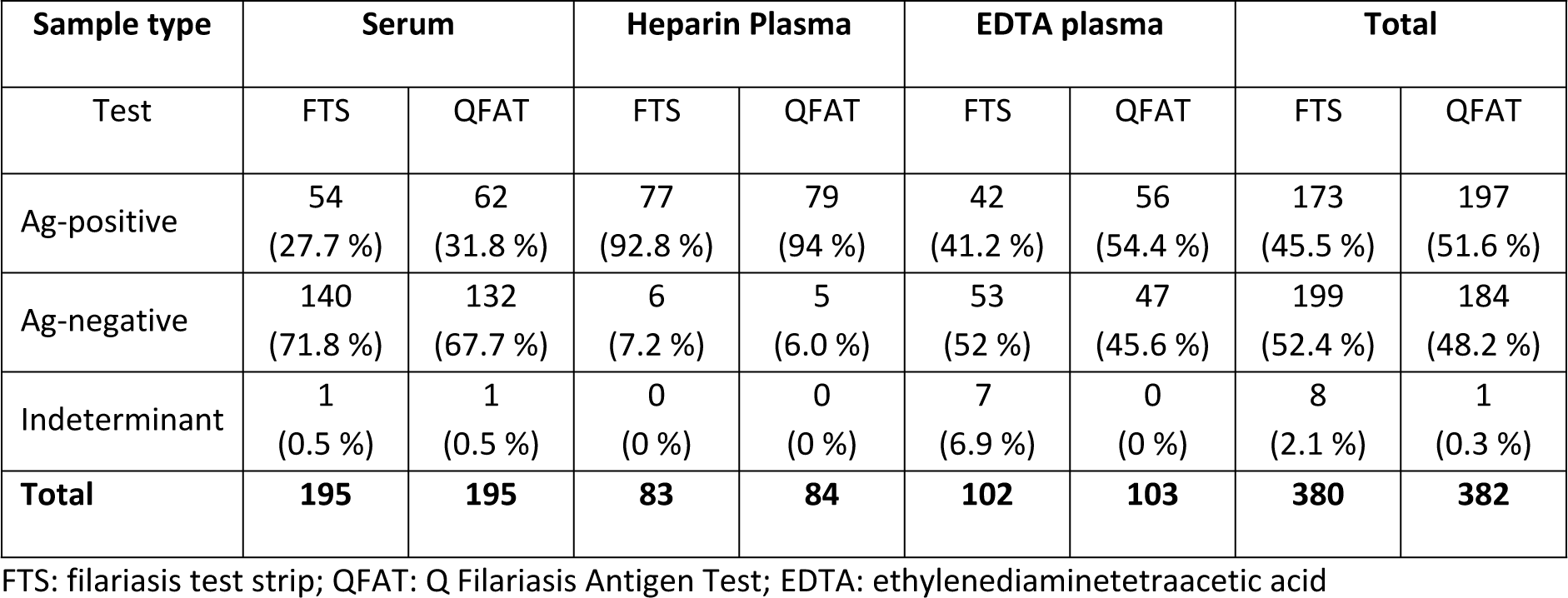
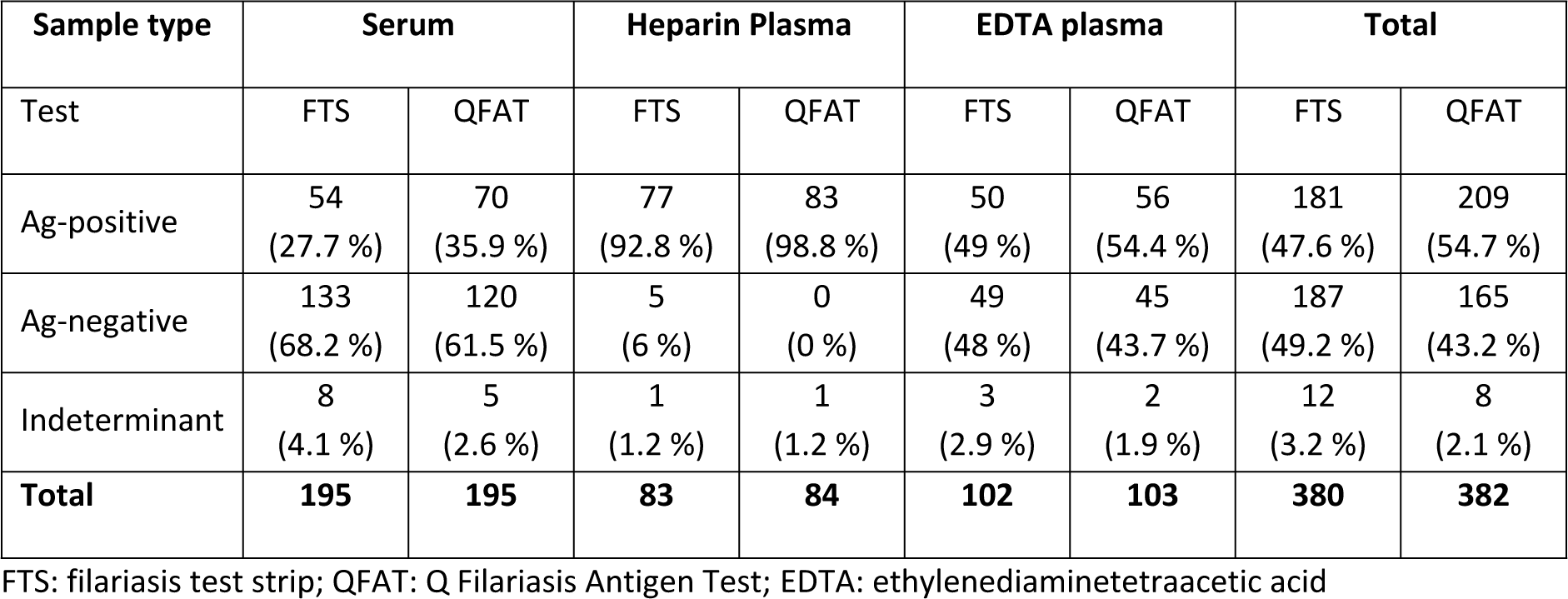
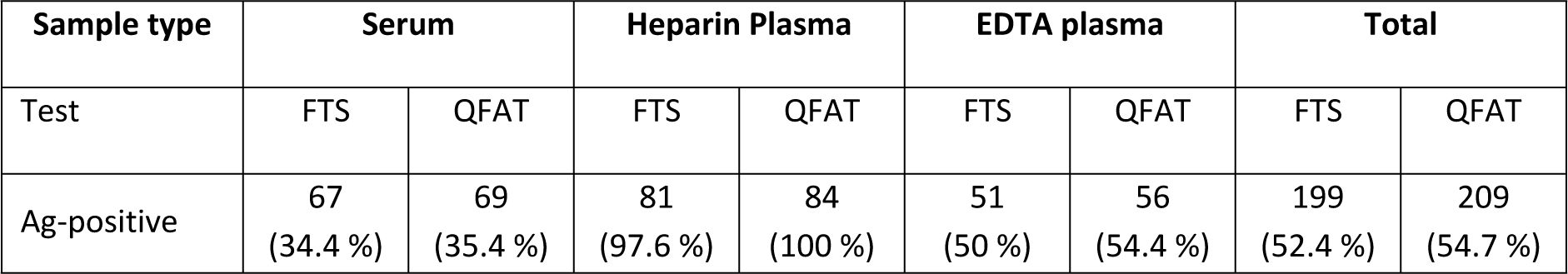
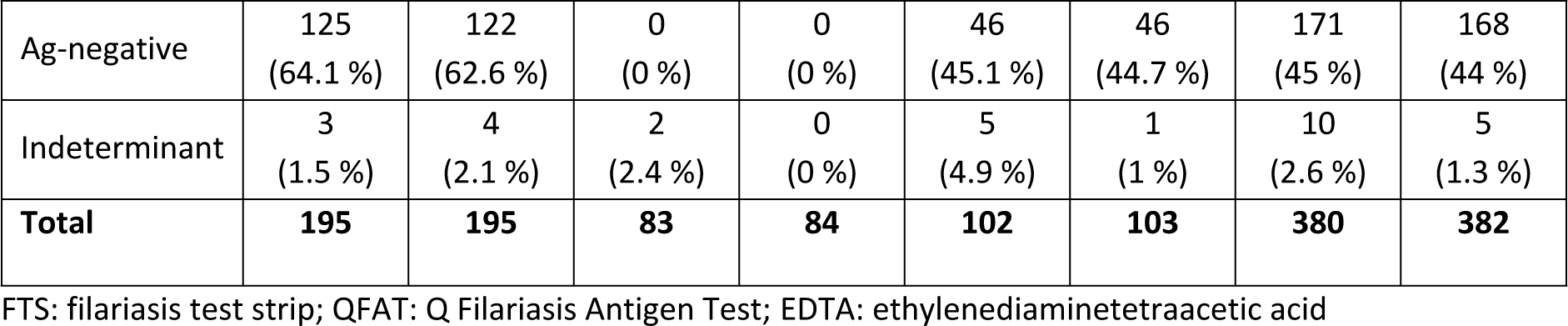
A: Overall antigen (Ag) positivity as determined by FTS and QFAT in this study in samples from LF endemic areas at the 10-minute reading B: Overall antigen (Ag) positivity as determined by FTS and QFAT in this study in samples from LF endemic areas at the one-hour reading. C: Overall antigen (Ag) positivity as determined by FTS and QFAT in this study in samples from LF endemic areas at the 24-hours reading.

| Sample type | Serum |  | Heparin Plasma |  | EDTA plasma |  | Total |  |
| --- | --- | --- | --- | --- | --- | --- | --- | --- |
|  | FTS | QFAT | FTS | QFAT | FTS | QFAT | FTS | QFAT |
| Ag-positive | 54<br>(27.7 %) | 62<br>(31.8 %) | 77<br>(92.8 %) | 79<br>(94 %) | 42<br>(41.2 %) | 56<br>(54.4 %) | 173<br>(45.5 %) | 197<br>(51.6 %) |
| Ag-negative | 140<br>(71.8 %) | 132<br>(67.7 %) | 6<br>(7.2 %) | 5<br>(6.0 %) | 53<br>(52 %) | 47<br>(45.6 %) | 199<br>(52.4 %) | 184<br>(48.2 %) |
| Indeterminant | 1<br>(0.5 %) | 1<br>(0.5 %) | 0<br>(0 %) | 0<br>(0 %) | 7<br>(6.9 %) | 0<br>(0 %) | 8<br>(2.1 %) | 1<br>(0.3 %) |
| **Total** | **195** | **195** | **83** | **84** | **102** | **103** | **380** | **382** |

FTS: filariasis test strip; QFAT: Q Filariasis Antigen Test; EDTA: ethylenediaminetetraacetic acid

| Sample type | Serum |  | Heparin Plasma |  | EDTA plasma |  | Total |  |
| --- | --- | --- | --- | --- | --- | --- | --- | --- |
|  | FTS | QFAT | FTS | QFAT | FTS | QFAT | FTS | QFAT |
| Ag-positive | 54<br>(27.7 %) | 70<br>(35.9 %) | 77<br>(92.8 %) | 83<br>(98.8 %) | 50<br>(49 %) | 56<br>(54.4 %) | 181<br>(47.6 %) | 209<br>(54.7 %) |
| Ag-negative | 133<br>(68.2 %) | 120<br>(61.5 %) | 5<br>(6 %) | 0<br>(0 %) | 49<br>(48 %) | 45<br>(43.7 %) | 187<br>(49.2 %) | 165<br>(43.2 %) |
| Indeterminant | 8<br>(4.1 %) | 5<br>(2.6 %) | 1<br>(1.2 %) | 1<br>(1.2 %) | 3<br>(2.9 %) | 2<br>(1.9 %) | 12<br>(3.2 %) | 8<br>(2.1 %) |
| **Total** | **195** | **195** | **83** | **84** | **102** | **103** | **380** | **382** |

FTS: filariasis test strip; QFAT: Q Filariasis Antigen Test; EDTA: ethylenediaminetetraacetic acid

| Sample type | Serum |  | Heparin Plasma |  | EDTA plasma |  | Total |  |
| --- | --- | --- | --- | --- | --- | --- | --- | --- |
|  | FTS | QFAT | FTS | QFAT | FTS | QFAT | FTS | QFAT |
| Ag-positive | 67<br>(34.4 %) | 69<br>(35.4 %) | 81<br>(97.6 %) | 84<br>(100 %) | 51<br>(50 %) | 56<br>(54.4 %) | 199<br>(52.4 %) | 209<br>(54.7 %) |
| Ag-negative | 125<br>(64.1 %) | 122<br>(62.6 %) | 0<br>(0 %) | 0<br>(0 %) | 46<br>(45.1 %) | 46<br>(44.7 %) | 171<br>(45 %) | 168<br>(44 %) |
| Indeterminant | 3<br>(1.5 %) | 4<br>(2.1 %) | 2<br>(2.4 %) | 0<br>(0 %) | 5<br>(4.9 %) | 1<br>(1 %) | 10<br>(2.6 %) | 5<br>(1.3 %) |
| **Total** | **195** | **195** | **83** | **84** | **102** | **103** | **380** | **382** |

FTS: filariasis test strip; QFAT: Q Filariasis Antigen Test; EDTA: ethylenediaminetetraacetic acid

Of note, Table 1A shows a higher number of indeterminant results for FTS with EDTA plasma while there were no indeterminant results for QFAT with this sample type. Tables 1B and 1C show the equivalent numbers for the 1-hour and 24-hour readings, respectively.

### Concordance between tests

There was a high level of concordance between tests, as shown in Table 2. Concordance increased over time, with the lowest level of concordance occurring at 10 mins (93.5%) and the highest recorded at 24 hours (98.8%). Kappa values followed the same pattern, with Kappa (K) = 0.88 (95% CI 0.83 to 0.92) at 10 minutes, and a K value of 0.98 (95% CI 0.95 to 1) occurring at 24 hours (Table 2). Overall, the Kappa agreement statistic suggested “excellent” agreement between the two tests.

**Table 2.** Concordance between tests, excluding indeterminant results, for all timepoints.

| Time-points | FTS | QFAT |  | N | Concordance (%) | Kappa (95% CI) |
| --- | --- | --- | --- | --- | --- | --- |
|  |  | Positive | Negative |  |  |  |
| 10 minutes | Positive | 167 | 6 | 417 | 93.5 | 0.87<br>(0.82-0.92) |
|  | Negative | 21 | 223 |  |  |  |
| 1 hour | Positive | 180 | 1 | 407 | 94.8 | 0.90<br>(0.85-0.94) |
|  | Negative | 20 | 206 |  |  |  |
| 24 hours | Positive | 196 | 1 | 412 | 98.8 | 0.98<br>(0.95-1.0) |
|  | Negative | 4 | 211 |  |  |  |

CI: Confidence interval; FTS: Filariasis Test Strip; QFAT: Q Filariasis Antigen Test

### Test performance compared with prior antigen or antigen/antibody results

At 10 minutes, when comparing predetermined Ag results, the sensitivity and negative predictive value of the FTS was lower than for QFAT. The specificity and positive predictive value at this timepoint for FTS and QFAT were comparable (Table 3). When comparing results of previously determined ‘Ag or any Ab’ results, the sensitivity and negative predictive values were again lower for both FTS and QFAT. However, sensitivity and negative predictive values were higher for QFAT than FTS.

**Table 3.** Sensitivity, specificity, positive predictive value and negative predictive value of FTS and QFAT at the 10-minute reading, compared to prior antigen and ‘antigen or any antibody’ test result.

| Test | Positive for antigen only |  | Positive for antigen or any antibody |  |
| --- | --- | --- | --- | --- |
| Biomarker comparison | FTS (n=417) | QFAT (n=427) | FTS (n=417) | QFAT (n=427) |
| **Sensitivity** | 86% | 92% | 64% | 70% |
| **% (95% CI)** | (80-90%) | (88-96%) | (58-70%) | (65-76%) |
| **Specificity** | 99% | 98% | 99% | 99% |
| **% (95% CI)** | (97-100%) | (95-99%) | (96-100%) | (95-100%) |
| **PPV (95% CI)** | 0.99<br>(0.96-1.00) | 0.98<br>(0.95-0.99) | 0.99<br>(0.97-1.00) | 0.99<br>(0.96-1.00) |
| **NPV (95% CI)** | 0.88<br>(0.83-0.92) | 0.93<br>(0.89-0.96) | 0.61<br>(0.54-0.67) | 0.64<br>(0.58-0.71) |

Positive predictive values (PPV); Negative predictive values (NPV); CI: confidence interval; FTS: Filariasis Test
Strip; QFAT: Q Filariasis Antigen Test

### Concordance and performance of tests with EDTA plasma samples only

Since questions have been raised by users about the suitability of FTS for EDTA plasma, we investigated the performance of the tests specifically for those samples. Concordance and sensitivity/specificity are shown in Supplementary Table 3. Most of the discordant samples between tests were of EDTA plasma sample type, for which QFAT appears more accurate than FTS.

### Change in test performance over time

Some test results changed between readings over time, mainly from negative to positive (Supplementary Table 4). For FTS, four negatives at 10 minutes became positive at 1 hour, while a further 18 became positive at 24 hours (Supplementary Table 4a). Positives at 10 minutes generally remained stable; only one became indeterminant at 1 hour but reverted to positive at 24 hours. For QFAT, 10 negatives at 10 minutes became positive at 1 hour. Of these, one positive reverted to negative at 24 hours and one became indeterminant. A further two samples changed from negative to positive at 24 hours (Supplementary Table 4b).

Test performance increased over time. Figure 2a shows that FTS sensitivity increased from 86% at the 10-minute reading to 89% after an hour, reaching a final value of 97% after 24 hours. Similarly, QFAT sensitivity was at its lowest at the 10-minute reading (89%) and increased to its highest values at the 1 hour and 24 hours readings. Thus, the difference in sensitivity and specificity between FTS and QFAT decreased over time. Corresponding results for positive and negative predictive values are given in Figure 2b.

**Fig 2.**
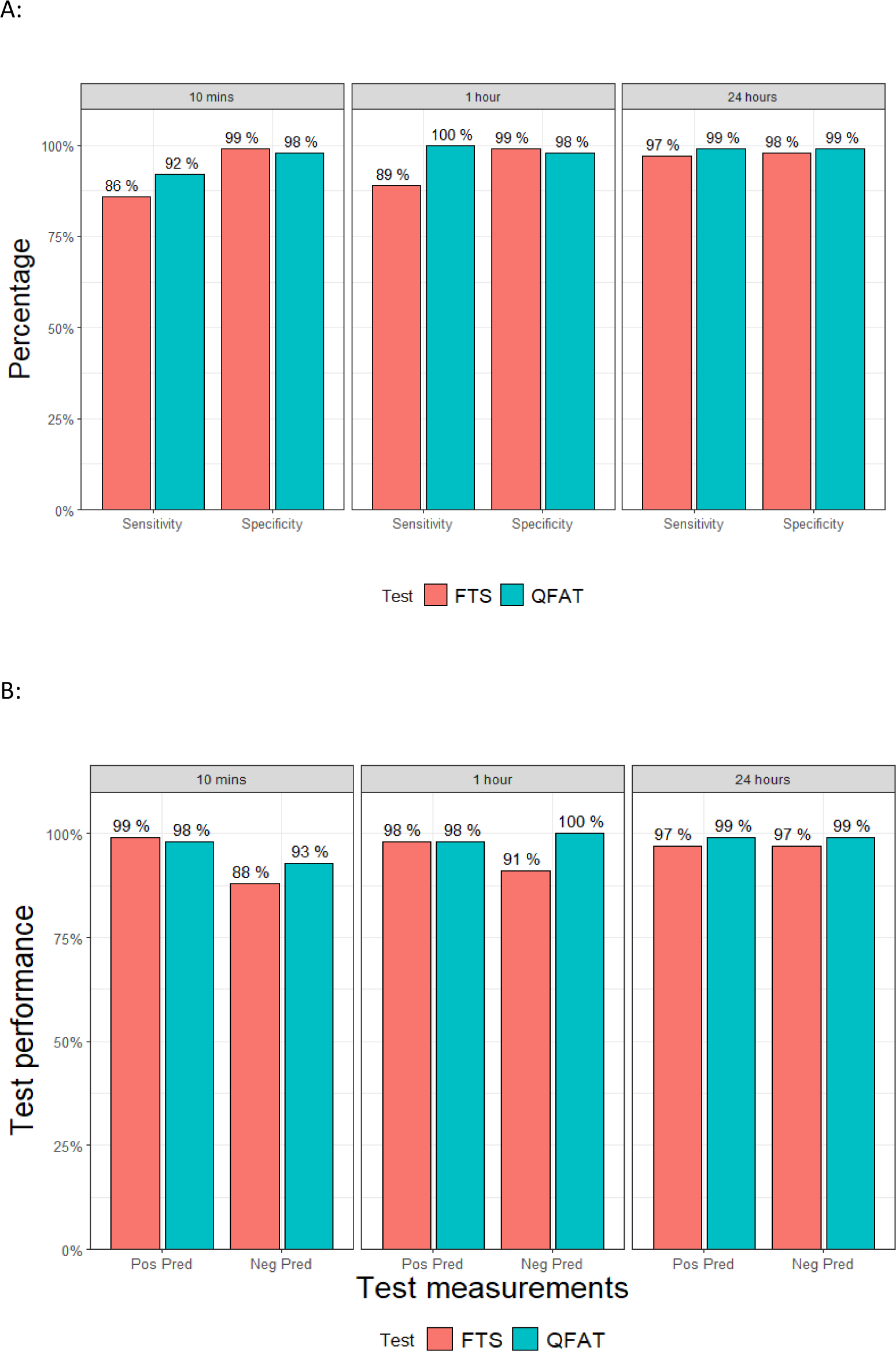
a. Sensitivity and specificity of FTS and QFAT, compared to prior Ag results, at three time points. b. Positive and negative predictive values of FTS and QFAT, compared to prior Ag results, at three time points.

### Performance in Mf-positive samples

A total of 69 samples were previously positive for Mf. After removal of one missing QFAT test, the remaining 68 samples all had positive QFAT results at 10 minutes. For FTS, after removal of one missing sample and two indeterminant results, FTS missed three of 66 (4.6%) Mf-positive samples at the 10-minute reading. Of note, two of the three false negative samples were from the same person (serum and heparin plasma samples).

### Cross-reactions with other helminth infections

Of the 20 *Strongyloides-*positive samples from humans, 19 tested with QFAT and one tested with FTS were all Ag-negative. Samples from four dogs infected with *Acanthochilonema reconditum* (1), *Cercopithifilaria bainae* (2), and *Dirofilaria repens* (1) were negative by both tests, while samples from one dog with *D. repens* and one with *Onchocerca lupi* were positive by both tests.

### Intensity score over time, by test type and observer

The proportion of intensity score values assigned by observers was different between FTS and QFAT. Supplementary Figure 2 shows that both observers reported a larger number of high intensity scores (test line denser than control line) using FTS than with QFAT. For the number of samples with results at time points included in each comparison, see Supplementary Fig 1.

In general, the line intensity score was less useful for QFAT than FTS because the stronger control line with QFAT means that almost all readings were classified as low or moderate intensity.

### Test acceptability

Two observers independently provided feedback on the tests. Both preferred QFAT over FTS for all aspects except the packaging, which was harder to open quickly for QFAT. Both mentioned that QFAT requires much less sample (10 or 20 uL vs 75 uL for FTS) and that QFAT was easier to use. QFAT requires an additional buffer step, which was mentioned as a disadvantage, but was clearer to read.

## Discussion

Our laboratory-based study found very high concordance between the FTS and QFAT results when read at 10 minutes. This study also found that QFAT had equivalent performance to FTS for detecting *W. bancrofti* Ag based on prior known results of LF antigen for samples from the Asia Pacific. Like FTS [22, 23, 11] (Dickson et al., 2018; Lau et al., 2020; Sheel et al., 2021), QFAT performed very well for detecting *W. bancrofti* circulating Ag in persons from endemic areas in Samoa, American Samoa, and Myanmar. No false positives were observed with non-exposed human sera.

Regarding test concordance in different sample types, we noted a higher number of positives with FTS than QFAT using EDTA plasma. False positives with FTS are a known reported issue with EDTA plasma among test users.

Compared with the main reference standard of previous Ag results, QFAT performed better than FTS in terms of sensitivity (92% for QFAT and 86% for FTS) at the 10-minute reading. Both tests were highly specific (≥98%). When compared to previous ‘Ag or any Ab’ results, sensitivity was lower for both tests (70% QFAT and 64% FTS) but specificity remained unchanged. This suggests that people who were Ab-positive but not Ag-positive do not constitute a pool of Ag positives in the population, undetected by either test.

The concordance between tests increased at 1 hour and 24 hours. Sensitivity of each test compared to prior results also increased over time, which suggests that the increase in concordance was real (i.e., tests were not accumulating false positives over time). These findings suggest that both tests were robust, and accuracy was not compromised even when reading time was unexpectedly delayed beyond the recommended time interval.

QFAT and FTS showed cross-reactivity with some dog samples positive for *O.lupi* and *D.repens*, but not *C. bainae* and *A. reconditum*. No cross-reactivity was seen for QFAT with *Strongyloides*-positive human samples. Further testing with other human and animal worm parasites is recommended. This study benefited from a controlled lab environment and a panel of previously well characterised samples. A limitation of this study was that the samples had been stored for long periods of time (several years at −70°C). Also, tests were done with the available samples of serum or plasma, including heparinized or EDTA anticoagulated plasma. The FTS test instructions are not explicit, but it is generally accepted that heparin rather than EDTA is the recommended anticoagulant for the test. Our results suggest that QFAT does not have this limitation and performs better than FTS for EDTA samples. In addition, selection of samples to include appropriate numbers of both serum and plasma meant that 74 individuals contributed two sample types. Previous Ag and Ab results on the samples had been determined by a variety of different methods (Og4C3 ELISA, ICT, FTS, Bm14 ELISA and Wb123 ELISA). Finally, the need for an additional buffer step for QFAT could be seen as a disadvantage, but opinions differ and some users regard lack of chase buffer for FTS as a disadvantage.

This laboratory evaluation shows that QFAT is a suitable rapid antigen test for use in GPELF and has advantages over FTS.

## Data Availability

The raw dataset has been lodged with TFGH and will be made available

## Abbreviations

Ag: Antigen
Ab: Antibody
LF: Lymphatic filariasis
QFAT: Q filariasis antigen test
FTS: Filariasis test strip
Mf: Microfilariae
WHO: World Health Organization
GPELF: Global Programme to Eliminate Lymphatic Filariasis
MDA: Mass drug administration
ICT: Immunochromatographic test
CI: Confidence interval
K: Cohen’s Kappa

## Acknowledgements

We are grateful to the following collaborators who provided blood samples and/or did previous testing: Jan Douglass, Khin Saw Aye, Ben Dickson, Sarah Sheridan, Meru Sheel, Take Naseri, Robert Thomsen, Saipale Fuimaono, Jesse Masson, Therese Kearns, Rogan Lee, and Alessio Giannelli. Jonathan King was the liaison between WHO NTD, PQ and Procurement departments and the manufacturer, without which the study would not have been possible. We thank Patrick Lammie (Task Force for Global Health) for supporting the study, FR3 for providing the positive control, WHO for providing FTS, and SD Biosensor for providing QFAT, and Julia S An for conducting training for QFAT over Zoom.

---

**Supplementary Table 1:** Profile and number of samples selected for the laboratory comparison of FTS and QFAT for LF antigen detection. The selected samples were collected from LF endemic areas, with known LF antigen and antibody status (using ICT, FTS or Og4C3 ELISA) determined prior to the current study.

| Country | American Samoa |  |  | Samoa |  | Myanmar | Total (% positive) |
| --- | --- | --- | --- | --- | --- | --- | --- |
| Year of sample collection | 2014 | 2016 | 2016 | 2019 | 2019 | 2014 |  |
| Sample type | SST Serum | SST Serum | Heparin Plasma | Heparin Plasma | EDTA Plasma | EDTA Plasma | All |
| **Antigen** |  |  |  |  |  |  |  |
| N with prior antigen results | 133 | 62 | 62 | 23 | 12 | 92 | 384 |
| Antigen-positive | 8 | 62 | 62 | 23 | 12 | 44 | 211 (54.9%) |
| Antigen-negative | 125 | NA | NA | NA | NA | 48 | 173 (45.1%) |
| **Antibodies** |  |  |  |  |  |  |  |
| Bm14 Ab-positive | 47 | 53 | 53 | NA | NA | 39 | 192 (55.0%) |
| Bm14 Ab-negative | 86 | 9 | 9 | NA | NA | 53 | 157 (45.0%) |
| Wb123 Ab-positive | 48 | 55 | 55 | NA | NA | 32 | 190 (54.4%) |
| Wb123 Ab-negative | 85 | 7 | 7 | NA | NA | 60 | 159 (45.6%) |
| Any antibody positive | 68 | 59 | 59 | NA | NA | 41 | 227 (65.0%) |
| Any antibody negative | 65 | 3 | 3 | NA | NA | 51 | 122 (35.1%) |
| **Antigen and/or antibody** |  |  |  |  |  |  |  |
| Antigen or any antibody positive | 69 | 62 | 62 | 23 | 12 | 51 | 279 (73.4%) |
| Antigen or any antibody negative | 64 | 0 | 0 | NA | NA | 41 | 101 (26.6%) |

EDTA: ethylenediaminetetraacetic acid; NA: not available

**Supplementary Table 2.** Number and percentage of results with each test that showed discordance between observers at 10 minutes.

|  | FTS |  | QFAT |  |  |
| --- | --- | --- | --- | --- | --- |
| Sample type | Total tested | Discordant results (%) | Total tested | Discordant results (%) | Chi-square (p-value) |
| **Endemic samples** |  |  |  |  |  |
| Total | 380 | 14 (3.7 %) | 382 | 7 (1.8 %) | 2.8 (0.096) |
| Serum | 195 | 6 (3.1 %) | 195 | 2 (1.0 %) | 1.5 (0.221) |
| Heparin Plasma | 83 | 1 (1.2 %) | 84 | 1 (1.2 %) | 0 (1.0) |
| EDTA Plasma | 102 | 7 (6.8 %) | 103 | 4 (3.9 %) | 0.8 (0.371) |
| **Non-endemic samples** |  |  |  |  |  |
| Total | 53 | 0 | 72 | 0 |  |
| Australian negative controls | 45 | 0 | 46 | 0 |  |
| Australian *Strongyloides*-positive | 1 | 0 | 19 | 0 |  |
| Dog serum | 7 | 0 | 7 | 0 |  |

FTS: filariasis test strip; QFAT: Q Filariasis Antigen Test; EDTA: ethylenediaminetetraacetic acid; p-value: probability under the assumption of no difference between tests (null hypothesis), of obtaining a result equal to or more extreme than what was actually observed.

**Supplementary Table 3.** A: Concordance for 10 min reading with EDTA plasma samples only. B: Sensitivity and specificity for 10 min reading with EDTA plasma samples only

| Reading time |  | QFAT-Positive | QFAT-Negative | Total | Concordance (%) | Kappa (95% CI) |
| --- | --- | --- | --- | --- | --- | --- |
| 10 mins | FTS-Positive | 41 | 1 | 42 |  |  |
|  | FTS-Negative | 7 | 46 | 53 |  |  |
|  | Total | 48 | 47 | 95 | 91.6 | 0.83 (0.72-0.94) |

| Reading time | Test | Sensitivity | Specificity | N |
| --- | --- | --- | --- | --- |
| 10 min | FTS | 85% (72% to 94%) | 96% (86% to 99%) | 95 |
|  | QFAT | 96% (87% to 100%) | 94% (83% to 99%) | 103 |

**Supplementary Table 4.** Samples with changes over time in reactivity for a) FTS and b) QFAT at 10 mins, 1 hour and 24 hours.

| **ID** | **Country** | **Sample type** | **FTS result at**<br>**10 minutes** | **FTS result at**<br>**1 hour** | **FTS result at**<br>**24 hours** |
| --- | --- | --- | --- | --- | --- |
| 296 | Myanmar | EDTA Plasma | Indeterminate | Indeterminate | Positive |
| 370 | Myanmar | EDTA Plasma | Indeterminate | Positive | Indeterminate |
| 271 | Samoa | EDTA Plasma | Indeterminate | Positive | Positive |
| 305 | Myanmar | EDTA Plasma | Indeterminate | Positive | Positive |
| 327 | Myanmar | EDTA Plasma | Indeterminate | Positive | Positive |
| 341 | Myanmar | EDTA Plasma | Indeterminate | Positive | Positive |
| 351 | Myanmar | EDTA Plasma | Indeterminate | Positive | Positive |
| 328 | Myanmar | EDTA Plasma | Negative | Indeterminate | Negative |
| 335 | Myanmar | EDTA Plasma | Negative | Indeterminate | Negative |
| 302 | Myanmar | EDTA Plasma | Negative | Negative | Indeterminate |
| 356 | Myanmar | EDTA Plasma | Negative | Negative | Indeterminate |
| 361 | Myanmar | EDTA Plasma | Negative | Negative | Indeterminate |
| 309 | Myanmar | EDTA Plasma | Negative | Negative | Positive |
| 354 | Myanmar | EDTA Plasma | Negative | Negative | Positive |
| 364 | Myanmar | EDTA Plasma | Negative | Positive | Indeterminate |
| 331 | Myanmar | EDTA Plasma | Negative | Positive | Positive |
| 86 | Am Samoa | HEP Plasma | Negative | Negative | Indeterminate |
| 103 | Am Samoa | HEP Plasma | Negative | Negative | Indeterminate |
| 70 | Am Samoa | HEP Plasma | Negative | Negative | Positive |
| 88 | Am Samoa | HEP Plasma | Negative | Negative | Positive |
| 90 | Am Samoa | HEP Plasma | Negative | Negative | Positive |
| 268 | Samoa | HEP Plasma | Negative | Positive | Positive |
| 133 | Am Samoa | Serum | Negative | Indeterminate | Negative |
| 147 | Am Samoa | Serum | Negative | Indeterminate | Negative |
| 151 | Am Samoa | Serum | Negative | Indeterminate | Negative |
| 12 | Am Samoa | Serum | Negative | Indeterminate | Positive |
| 62 | Am Samoa | Serum | Negative | Indeterminate | Positive |
| 128 | Am Samoa | Serum | Negative | Indeterminate | Positive |
| 24 | Am Samoa | Serum | Negative | Negative | Indeterminate |
| 170 | Am Samoa | Serum | Negative | Negative | Indeterminate |
| 7 | Am Samoa | Serum | Negative | Negative | Positive |
| 26 | Am Samoa | Serum | Negative | Negative | Positive |
| 28 | Am Samoa | Serum | Negative | Negative | Positive |
| 43 | Am Samoa | Serum | Negative | Negative | Positive |
| 57 | Am Samoa | Serum | Negative | Negative | Positive |
| 59 | Am Samoa | Serum | Negative | Negative | Positive |
| 129 | Am Samoa | Serum | Negative | Negative | Positive |
| 157 | Am Samoa | Serum | Negative | Negative | Positive |
| 169 | Am Samoa | Serum | Negative | Negative | Positive |
| 130 | Am Samoa | Serum | Negative | Positive | Positive |
| 31 | Am Samoa | Serum | Positive | Indeterminate | Positive |

| rowid | country | sampletype | QFAT result at<br>10 minutes | QFAT result at<br>1 hour | QFAT result at<br>24 hours |
| --- | --- | --- | --- | --- | --- |
| 334 | Myanmar | EDTA Plasma | Negative | Indeterminate | Indeterminate |
| 371 | Myanmar | EDTA Plasma | Negative | Indeterminate | Negative |
| 103 | Am Samoa | HEP Plasma | Negative | Indeterminate | Positive |
| 72 | Am Samoa | HEP Plasma | Negative | Positive | Positive |
| 74 | Am Samoa | HEP Plasma | Negative | Positive | Positive |
| 90 | Am Samoa | HEP Plasma | Negative | Positive | Positive |
| 169 | Am Samoa | Serum | Negative | Indeterminate | Indeterminate |
| 134 | Am Samoa | Serum | Negative | Indeterminate | Negative |
| 145 | Am Samoa | Serum | Negative | Indeterminate | Negative |
| 28 | Am Samoa | Serum | Negative | Indeterminate | Positive |
| 170 | Am Samoa | Serum | Negative | Negative | Indeterminate |
| 141 | Am Samoa | Serum | Negative | Positive | Negative |
| 41 | Am Samoa | Serum | Negative | Positive | Positive |
| 61 | Am Samoa | Serum | Negative | Positive | Positive |
| 62 | Am Samoa | Serum | Negative | Positive | Positive |
| 128 | Am Samoa | Serum | Negative | Positive | Positive |
| 130 | Am Samoa | Serum | Negative | Positive | Positive |
| 133 | Am Samoa | Serum | Negative | Positive | Positive |
| 157 | Am Samoa | Serum | Positive | Positive | Indeterminate |

**Supplementary Fig 1:**
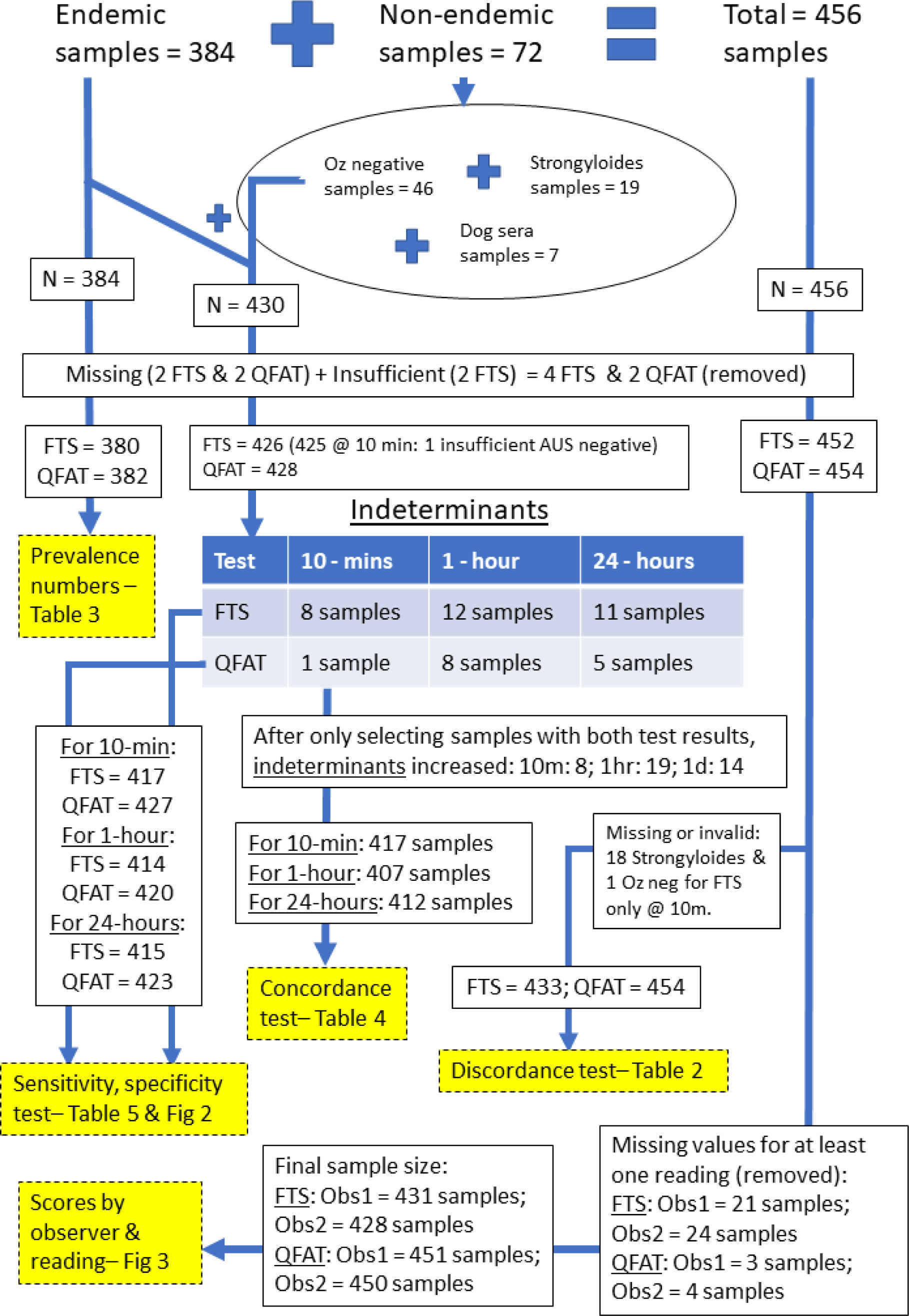
Flow chart of sample numbers in each comparison.

**Supplementary Fig 2.**
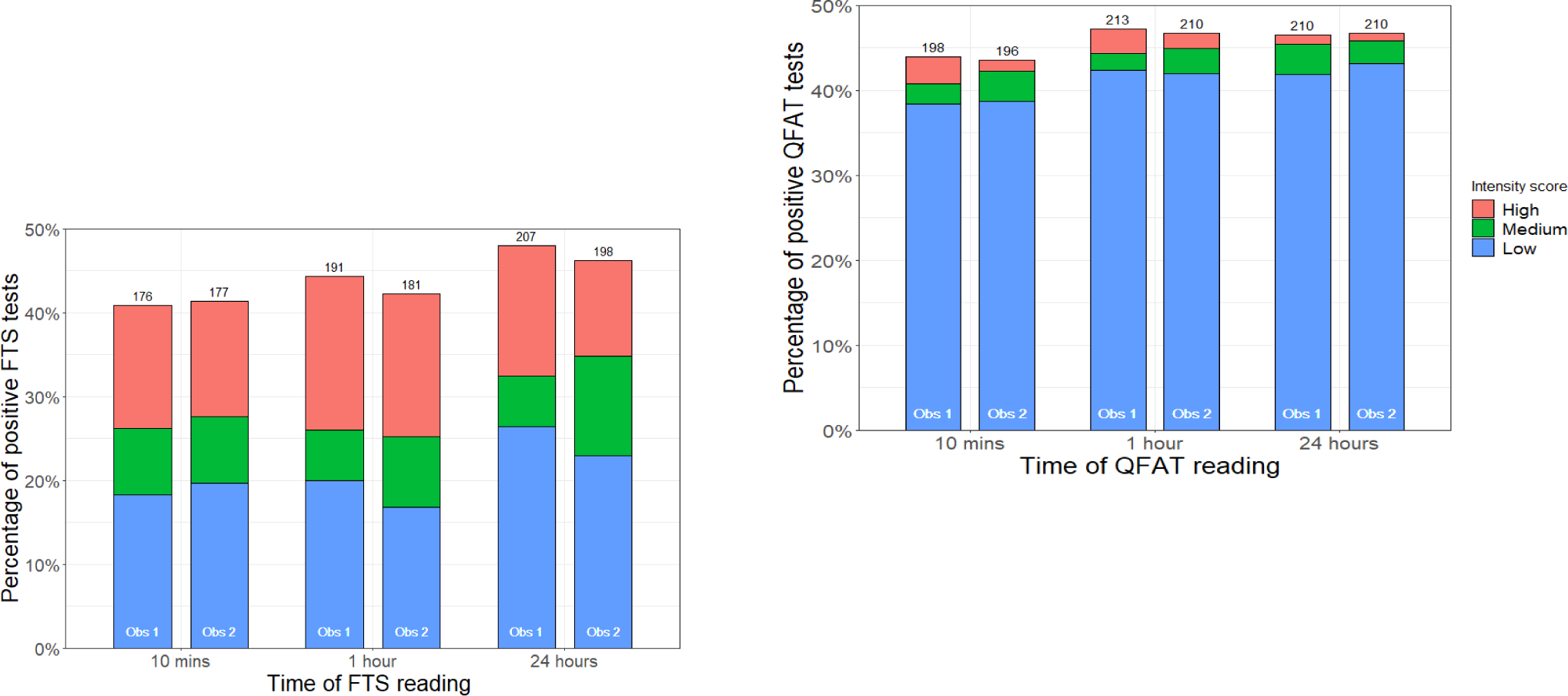
Test score by observer and time period, by test type.

